# Preferences for receiving study results among pregnant women participating in a phase III clinical trial in Papua New Guinea

**DOI:** 10.64898/2026.08.27.26361571

**Authors:** Alice Mengi, Mary Bagita-Vangana, Paula Tesine, Moses Laman, John W Bolnga, Maria Ome-Kaius, Janeth Kulimbao, Jacobeth Mase, Lennie S Mal, Hellen Mnjala, Grant Lee, Sarah A Cassidy-Seyoum, Kamala Thriemer, Holger W Unger

## Abstract

Disseminating study results to participants is an ethical responsibility for researchers but remains uncommon in low- and middle-income countries, and participants’ preferences for receiving study results are poorly understood. This study examined study result dissemination preferences among pregnant women in a phase III malaria prevention trial in Papua New Guinea (PNG). Participants completed an interviewer-administered questionnaire (survey) assessing their interest in and motivation for receiving trial results and preferences for dissemination methods and content. Associations between participants’ characteristics and dissemination preferences were explored using multivariable logistic regression analysis. Of 1172 trial participants, 96.0% (1125/1172) completed the survey, and of these 99.6% (1121/1125) wanted to learn about the trial results. The main motivation factors driving participants’ interest were an acknowledgment of their contribution to research (51.7%; n=579) and a better understanding of the study (45.0%; n=505). Most participants (78.9%; n=884) wanted to learn about the trial findings through written summary and a group meeting with other participants at the nearest clinic (31.1%, n=349). Multivariable regression analysis indicated that participants from rural/peri-urban clinics were more likely to choose non-electronic media dissemination approaches such as a group meeting as compared to urban-dwelling participants. Frequently selected items (>50% of participants) for content included information regarding ‘good results of the study,’ ‘purpose of the study,’ ‘medical treatment advances,’ ‘results specific to me,’ and ‘how study was conducted.’ There was heterogenicity in the preference for dissemination content: compared to urban clinics rural clinics are less likely to want to learn about ‘how and why study was conducted’ and ‘medical and scientific advances.’ Overall, the majority wanted to learn about trial results, highlighting the importance of integrating dissemination into research activities in PNG. Variation in preferences for mode and content of dissemination between study clinics suggests that dissemination activities could be tailored to local context and preferences.

## Background

Community engagement is increasingly recognised as an important component of ethical and effective clinical research (1, 2). It involves collaboration between community members, researchers and other stakeholders to address health-related issues and improve health outcomes. In clinical research, community engagement contributes to identification and recruitment of eligible participants, supports retention and follow-up, helps to address rumours or misconceptions, and can facilitate rapid responses during public health emergencies (3). However, community engagement is increasingly viewed not only as a strategy to facilitate trial implementation, but also as an ethical imperative grounded in principles of respect, reciprocity, transparency, and accountability to participating communities (1). Community engagement aims to build trust and mutual understanding between researchers and communities and to ensure that research is conducted in ways that are responsive to community needs and expectations (4).

An important, but often underemphasised, component of community engagement is the dissemination of aggregate research findings to trial participants and communities following study conclusion (2, 5, 6). Traditionally, dissemination has focused on academic and policy audiences through peer-reviewed publications, conference presentations, stakeholder meetings, websites, and media platforms (7). While these approaches are important for translating findings into policy and practice, they do not necessarily ensure that participants themselves receive the results of studies in which they took part.

Evidence suggests that research participants generally want to receive study results, with most studies reporting that a high proportion of participants want to learn about the outcomes of the research they participated, although the level of interest varies between settings (8, 9). Dissemination of study findings to participants may promote transparency, strengthen trust in researchers, improve understanding of health and research, and support future community engagement in research activities (5, 10). However, relatively little is known about how participants prefer to receive study results, particularly in low- and middle-income countries where dissemination activities are rarely evaluated and reported (5). Existing evidence is heavily concentrated in high-income settings and may not be directly transferable to other contexts (5). The limited available evidence suggests that dissemination preferences can vary substantially between populations and settings. For example, a recent multi-country study found that participants in Pakistan preferred summaries in their own language, those in Cambodia preferred a letter or phone call explaining the results, whereas participants in Ethiopia preferred a community meeting to learn about study results (11). In the same study, preferences also differed in regard to the type of information participants were interested in, with many expressing interests in learning about the study purpose, as opposed to overall study findings, or broader scientific implications (11). Reasons for why dissemination of research results to study participants is rarely done in low-resource settings may include limited funding, insufficient expertise, logistical challenges, and limited community engagement activities during trial planning and implementation (5). In rural and remote settings, dissemination may be further complicated by structural barriers such as poor road conditions, unreliable electricity supply, limited mobile and internet connectivity, and social instability such as conflict (12).

Papua New Guinea (PNG), a lower-middle income country in the Western Pacific region, hosts numerous clinical research studies, many of which include participants from rural and remote settings (13–15). To build further evidence on how dissemination should be conducted in the context of low- and middle-income countries, an evaluation of participants’ preferences was included as part of a malaria in pregnancy prevention clinical trial conducted in Madang Province, PNG (16). The objective of this study was to determine study participants’ preferences regarding the conduct and content of result dissemination.

## Methods

### Study context

This cross-sectional questionnaire-based survey was undertaken amongst pregnant women participating in the SAPOT trial, which evaluated intermittent preventative treatment in pregnancy (IPTp) with sulphadoxine-pyrimethamine plus dihydroartemisinin-piperaquine for the prevention of malaria infection and adverse pregnancy outcomes (NCT05426434). The trial was conducted across five clinical sites in Madang Province, PNG: Town Clinic, Madang Provincial Hospital, Yagaum Rural Hospital, Alexishafen Health Centre and Mugil Health Centre. The trial commenced enrolment on August 31, 2022, and follow-up concluded on the March 24, 2025.(16)

### Data collection

A questionnaire assessing dissemination preferences was nested in trial case record forms (Supplemental table 1). The questionnaire was adapted from previous work and discussions with the study team.(5, 17) The survey was interviewer-administered in Tok Pisin, the lingua franca in Madang Province. The questionnaire assessed preferences around several outcome domains. These domains included a) interest in learning about findings of the trial (yes/no), b) reasons for wanting to receive results (participants allowed to select one of two response options), c) preferences for format and mode of dissemination (multiple selections possible), and d) preferences for content of dissemination information material (multiple selections possible). The questionnaire was administered at a single time point, during the first follow-up visit that participants attended after the trial enrolment visit, to minimise participant burden during enrolment (16). Completed case record forms were checked for accuracy and entered into a password-protected and cloud-based database (REDCap, Nashville, USA), and physical case record forms are stored in a locked cabinet at the PNG Institute of Medical Research (PNGIMR) office in Madang (18).

### Outcome variables

Across the domains for format and mode of dissemination and thematic content of dissemination, participants could choose more than one option. Individual questionnaire response items were subsequently combined and grouped into five broader dissemination preference categories to facilitate regression analyses (Supplemental table 2). For format and mode of dissemination, categories were ‘print paper media,’ ‘electronic media,’ ‘community-based group communication,’ ‘personalised individual communication,’ and ‘multiple preferences.’ Similarly, preferences for thematic content of the dissemination were grouped into five categories (Supplemental table 2). These were ‘how and why study was conducted,’ ‘medical and scientific advances on treatment,’ ‘potential for future for research and change in policies,’ ‘results specific to me,’ and ‘multiple preferences for content.’

### Predictor measures

A range of potential predictor variables collected as part of the trial were considered relevant for this analysis and included demographic, socioeconomic, behavioural, and clinical factors. Demographic predictor variables included study site (Alexishafen, Modilon/Town clinics, Mugil, Yagaum) as a proxy for residence, maternal age in years (categorised as 16-24, 25-34, ≥35), ethnicity (categorised as Madang versus non-Madang ethnicity), education level in years of formal schooling (categorised as 0-2 years, 3-8 years, 9-12 years, and tertiary education), marital status (categorised as married versus single/separated), and years of residence in study location (categorised as ≤2 years of residence versus >2 years of residence). Socioeconomic variables included maternal employment status (categorised as unemployed/student, informal, and formal) and cooking method (categorised as open fire/coals versus other). Behavioural factors included smoking (yes/no), chewing betelnut (yes/no), alcohol consumption (yes/no), and self-reported use of a mosquito net in the preceding night (yes/no). Clinical factors included gravidity (primigravida, 2-3 pregnancies and ≥4 pregnancies), gestational age at enrolment (≤20 versus >20 weeks), and gestational age at completion of the questionnaire (≤20 versus >20 weeks).

### Statistical analysis

The sample size of this study was pre-determined by the number of participants enrolled into parent trial. Demographic, socioeconomic, behavioural, and clinical characteristics of participants were categorised and summarised using frequencies and percentages.

Responses to the four questionnaire domains ‘interest in receiving study results,’ ‘reasons for wanting results,’ ‘preferences for format and mode,’ and ‘content’ were reported using frequencies and percentages.

Associations of demographic, socioeconomic, behavioural and clinical factors with the derived outcomes for ‘mode and format of dissemination’ and ‘content of dissemination information materials’ were subsequently explored. Univariable analysis was used to assess the association of each exposure variable with outcome measures of format and mode and content of dissemination materials, using the Pearson’s chi-square. Variables associated with at least one outcome for each domain at p<0.1 in univariable analysis were included in the multivariable logistic regression models. Stata 18.0 (StataCorp, College Station, TX, USA) was used for data analysis.

### Ethics

Ethical approvals were obtained from Menzies School of Health Research Ethics Committee (2021–4107), the Madang Provincial Health Authority Research Ethics Committee (04.21), the PNG Institute of Medical Research Institutional Review Board (2113), and the PNG Medical Research Advisory Committee (22.10). Written informed consent was obtained from all participants prior to enrolment into the trial. All participants confirmed that they agreed to partake in the dissemination preferences evaluation.

## Results

### Study population

A total of 1,172 participants were enrolled into the trial, and 1,125 (96.0%) completed a preference questionnaire; 47 participants did not complete a questionnaire because of loss of follow-up after trial enrolment. Almost all participants (n=1,121, 99.6%) for whom questionnaires were completed expressed an interest in learning about the trial results following study completion and analysis.

Of the 1,125 participants with questionnaire data, 820 (72.9%) were from rural (Mugil, Yagaum) and peri-urban clinics (Alexishafen) (Table 1). Most participants were aged 16-34 years (n=1,022, 90.8%), had enrolled into the trial after 20 gestational weeks (n=728, 64.7%), and fewer than one-fifth of participants had four or more pregnancies (n=222, 19.7%). Most participants had completed at least primary education (n=1,054, 93.8%), were formally employed or partook in informal income-generating activities (n=1,015, 90.2%) and were married (n=1,076, 95.6%). The majority of participants reported that they were currently chewing betel nut (n=993, 88.3%), while fewer reported smoking tobacco (n=306, 27.2%) or ever consuming alcohol (n=197, 17.5%) (Table 1).

**Table 1.**
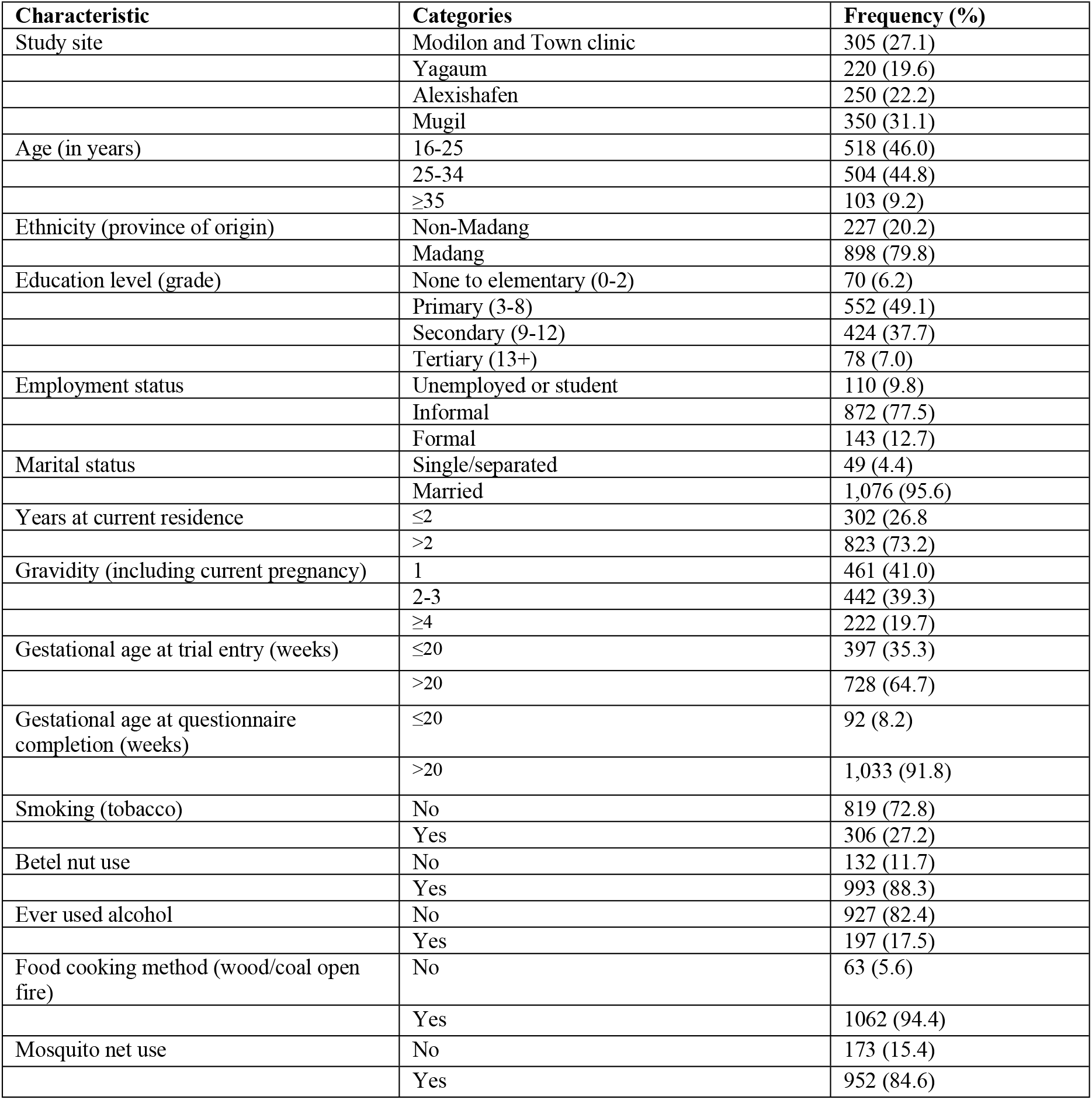
Demographic, socioeconomic, and clinical characteristics trial participants with questionnaire data (n=1,125)

| Characteristic | Categories | Frequency (%) |
| --- | --- | --- |
| Study site | Modilon and Town clinic | 305 (27.1) |
|  | Yagaum | 220 (19.6) |
|  | Alexishafen | 250 (22.2) |
|  | Mugil | 350 (31.1) |
| Age (in years) | 16-25 | 518 (46.0) |
|  | 25-34 | 504 (44.8) |
|  | ≥35 | 103 (9.2) |
| Ethnicity (province of origin) | Non-Madang | 227 (20.2) |
|  | Madang | 898 (79.8) |
| Education level (grade) | None to elementary (0-2) | 70 (6.2) |
|  | Primary (3-8) | 552 (49.1) |
|  | Secondary (9-12) | 424 (37.7) |
|  | Tertiary (13+) | 78 (7.0) |
| Employment status | Unemployed or student | 110 (9.8) |
|  | Informal | 872 (77.5) |
|  | Formal | 143 (12.7) |
| Marital status | Single/separated | 49 (4.4) |
|  | Married | 1,076 (95.6) |
| Years at current residence | ≤2 | 302 (26.8) |
|  | >2 | 823 (73.2) |
| Gravidity (including current pregnancy) | 1 | 461 (41.0) |
|  | 2-3 | 442 (39.3) |
|  | ≥4 | 222 (19.7) |
| Gestational age at trial entry (weeks) | ≤20 | 397 (35.3) |
|  | >20 | 728 (64.7) |
| Gestational age at questionnaire completion (weeks) | ≤20 | 92 (8.2) |
|  | >20 | 1,033 (91.8) |
| Smoking (tobacco) | No | 819 (72.8) |
|  | Yes | 306 (27.2) |
| Betel nut use | No | 132 (11.7) |
|  | Yes | 993 (88.3) |
| Ever used alcohol | No | 927 (82.4) |
|  | Yes | 197 (17.5) |
| Food cooking method (wood/coal open fire) | No | 63 (5.6) |
|  | Yes | 1062 (94.4) |
| Mosquito net use | No | 173 (15.4) |
|  | Yes | 952 (84.6) |

### Primary motivation for wanting to hear about study results

Participants who expressed an interest in receiving study results were subsequently asked about their motivation for wanting to receive the findings. More than half of the participants (51.7%, 579/1121) reported that receiving study results would acknowledge their contribution to research, while 45.0% (505/1121) wished to better understand the study in an easy and accessible way; 37 participants (3.3%, 37/1121) who initially wanted to receive study results did not provide a response.

### Preferences for format and mode of dissemination of the study results

Participant preferences for format and mode of dissemination are illustrated in Figure 1. The most commonly requested format for receiving results was a letter in Tok Pisin (the local lingua franca) or English (78.9%, n=884/1121), followed by a meeting with other participants at the nearest clinic (31.1%, n=349/1121). A phone call (14.5%, n=162/1121) and a short message service or text message (14.2%, n=159/1121) were selected less frequently. The least preferred methods were electronic formats including WhatsApp/messenger (2.0%, n=22/1121), email (1.1%, n=14/1121), and results presented on a website (0.1%, n=1/1121) or on Twitter/X (n=0/1121). Two participants did not provide a response to this questionnaire item.

**Figure 1.**
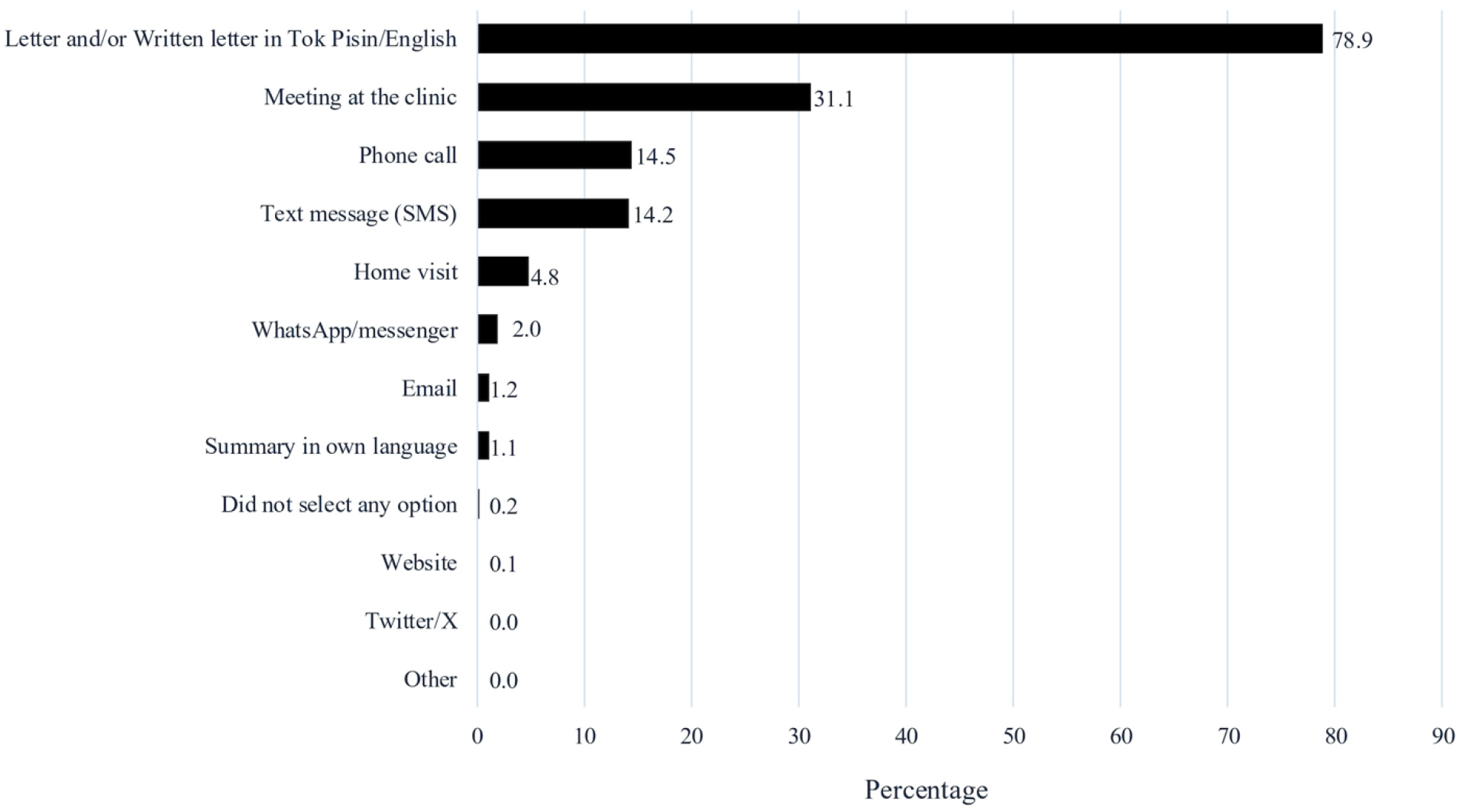
Participant preferences for the format and mode of dissemination of study results. Participants could select more than one option (n=1,121).

### Factors associated with preference for format and mode of dissemination

In univariable analyses, study site, ethnicity, education level, employment status, years of residence, gravidity, gestational age at questionnaire completion, betel nut use, alcohol use, cooking method, and mosquito net use were associated with at least one dissemination preference outcome (p<0.10) (Supplementary Table 3). These were subsequently included as covariates in multivariable models. Study site emerged as the strongest and most consistent predictor of dissemination preferences across all five outcomes (Table 2). Study site and the gestational age at questionnaire were the only predictors associated with all five outcomes. Years of residence, gestational age at trial entry, gravidity, betel nut use, alcohol use, cooking methods, and mosquito net use were not associated with content preferences (Table 2). In the multivariable analyses, study site, ethnicity, employment status, and gestational age at questionnaire remained associated with at least one content preference outcome (Table 2).

**Table 2:**
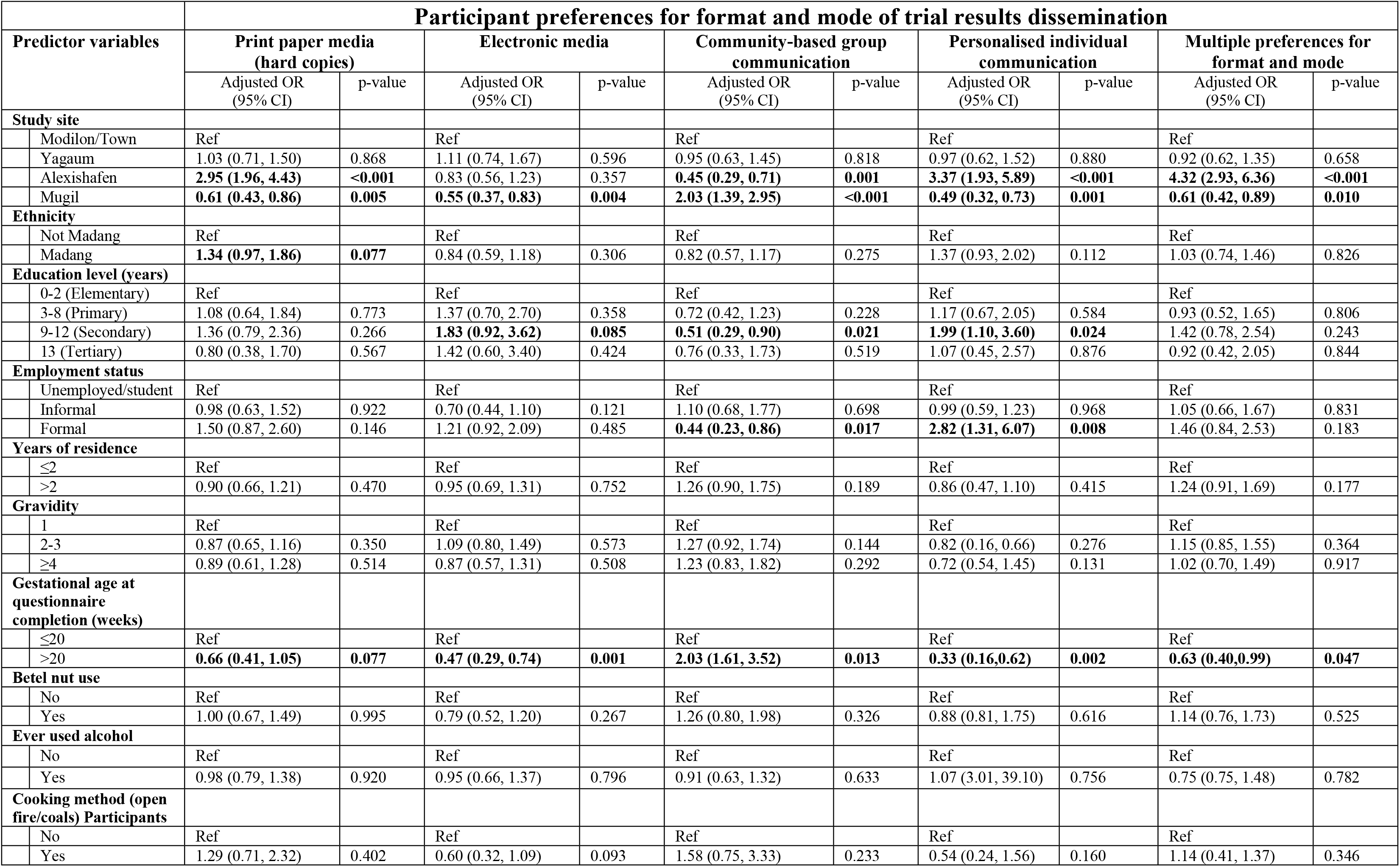
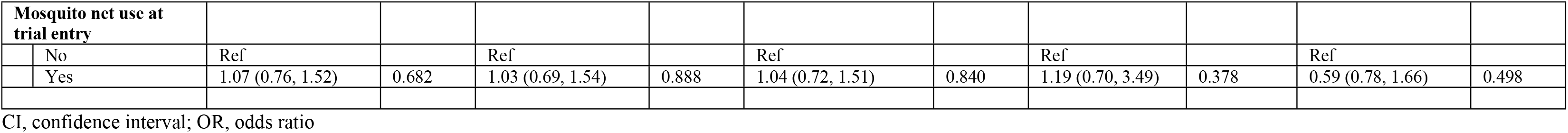
Multivariable analysis – participant preferences for format and mode of trial results dissemination (n=1,119)

#### Print media

In the multivariable analysis, study site was the only factor associated with a preference for print based dissemination (Table 2). Compared with participants attending the urban clinics (Modilon/Town Clinic), women attending the peri-urban (Alexishafen) clinic were more likely to prefer print media (adjusted odds ratio [aOR] 2.95, 95% confidence interval [CI] 1.96, 4.43), whereas women attending Mugil, the most rural clinic site, were less likely to do so (aOR 0.61%, 95%CI: 0.43, 0.86).

#### Electronic media

Study site and gestational age at questionnaire completion were associated with preference for electronic media formats in multivariable analysis (Table 2). Compared with women attending the urban Modilon/Town clinics, women attending Mugil were less likely to prefer electronic media (aOR 0.55, 95%CI 0.37, 0.83). Women completing the questionnaire after 20 weeks’ gestation were also less likely to prefer electronic dissemination (aOR 0.47, 95%CI 0.29, 0.74).

#### Community-based meetings

Study site, maternal education level, maternal employment status, and gestational age at questionnaire completion were associated with preferences for community-based group meetings in multivariable analysis (Table 2). Compared with women attending the urban Modilon/Town clinics, women attending Mugil were more likely to prefer community-based group meetings (aOR 2.03, 95%CI 1.39, 2.95), whereas women attending Alexishafen clinic were less likely to prefer this format (aOR 0.45, 95%CI 0.29, 0.71). Women who completed the questionnaire after 20 weeks’ gestation were also more likely to prefer community-based group meetings than those completing the questionnaire at ≤20 weeks’ gestation (aOR 2.03, 95%CI 1.61, 3.52). Compared to women with no formal or elementary education, those with secondary education (aOR 0.51, 95 %CI 0.29, 0.90) were less likely to choose a group meeting to learn about study results. Similarly, women in formal employment were less likely to prefer a group meeting than unemployed women (aOR 0.44, 95%CI 0.23, 0.86).

#### Personalised communication

In multivariable analysis, study site, education level, maternal employment status, and gestational age at questionnaire completion remained associated with ‘personalised individual communication’ (Table 2). Compared with women attending the urban Modilon/Town clinics, women attending Alexishafen were more likely to prefer personalised individual communication (aOR 3.37, 95%CI 1.93, 5.89). Women with secondary education were also more likely to prefer personalised communication than women with no formal or elementary education (aOR 1.99, 95%CI 1.0, 3.60), as were women in formal employment compared with unemployed women (aOR 2.82, 95%CI 1.31, 6.07). Women completing the questionnaire after 20 weeks’ gestation were less likely to prefer personalised individual communication than those completing the questionnaire at ≤20 weeks’ gestation (aOR 0.33, 95%CI 0.16, 0.62).

#### Multiple preferences

Study site and gestational age at questionnaire completion were associated with choosing more than one format/mode of dissemination (Table 2). Compared with women attending the urban Modilon/Town clinics, women attending Alexishafen were more likely to prefer multiple dissemination strategies (aOR 4.32; 95%CI 2.93, 6.36) whereas women attending Mugil were less likely to do so (aOR 0.61, 95%CI 0.42, 0.89). Women completing the questionnaire after 20 weeks’ gestation were also less likely to prefer multiple dissemination formats than those completing the questionnaire at ≤20 weeks’ gestation (aOR 0.63, 95%CI 0.40, 0.99)

### Preferred content of dissemination material information

Participant preferences for content of dissemination information materials are illustrated in Figure 2. The most commonly requested content was positive study findings (66.2%, n=742/1121), followed by purpose of the study (62.5%, n=701/1121), advances in medical treatment (53.3%, n=598/1121), participants’ individual study results (50.3%, n=564/1121), and how the study was conducted (50.2%, n=563/1121). The least selected content included neutral study findings (13.1%, n=147/1121), potential policy changes arising from the study (23.4%, n=262/1121), and future research informed by the study results (25.2%, n=283/1121). An open-ended ‘Other’ option was selected by four participants but only one participant provided additional information, requesting information on how the study would protect mothers and infants. Three participants did not select any of the options.

**Figure 2:**
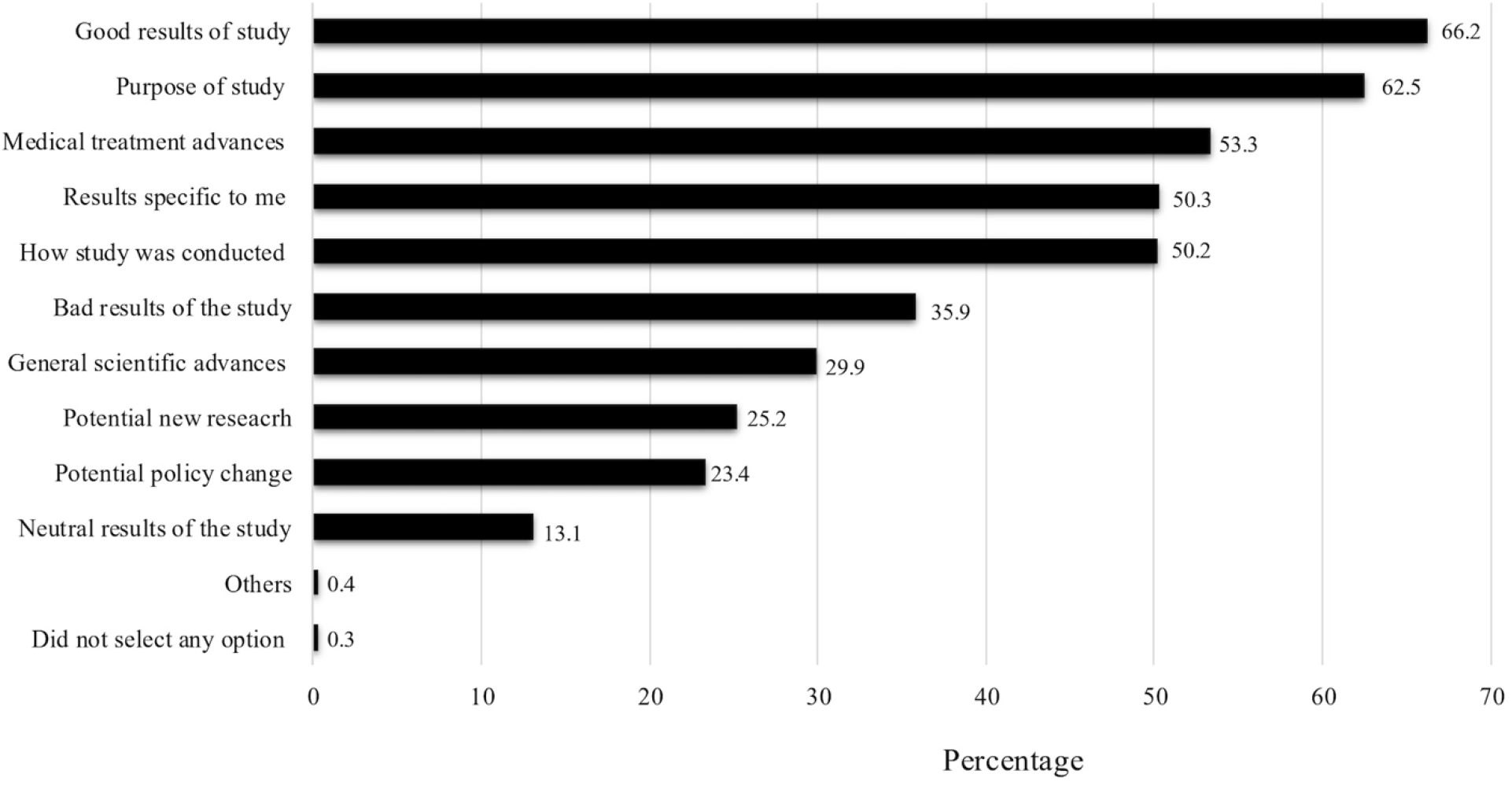
Participant preferences for content of dissemination of study results (n=1,121). Participants could select more than one response option.

### Key predictors for preferred content of dissemination information materials

On univariable analysis, study site, education level, employment status, gravidity, alcohol use, and cooking method were associated with at least one of the five content preference outcomes (Supplementary Table 4). Study site was the only predictor associated with all five outcomes. Maternal age, ethnicity, marital status, years of residence, gestational age at trial entry, gestational age at questionnaire completion, smoking, betel nut use, and mosquito net use were not associated with content preferences. In the multivariable analyses, study site, education level, employment status and gravidity remained associated with at least one content preference outcome (Table 3).

**Table 3:** Multivariable analysis – participant preferences for content of dissemination information materials (n=1,118)

| Participant preferences for content of dissemination information materials |  |  |  |  |  |  |  |  |  |  |  |
| --- | --- | --- | --- | --- | --- | --- | --- | --- | --- | --- | --- |
| Predictor variables |  | How and why study was conducted |  | Medical and scientific advances on treatment |  | Potential for future research and change in policies |  | Results specific to me |  | Multiple preferences for content |  |
|  |  | Adjusted OR (95% CI) | p-value | Adjusted OR (95% CI) | p-value | Adjusted OR (95% CI) | p-value | Adjusted OR (95% CI) | p-value | Adjusted OR (95% CI) | p-value |
| **Study site** |  |  |  |  |  |  |  |  |  |  |  |
|  | Modilon/Town | Ref |  | Ref |  | Ref |  | Ref |  | Ref |  |
|  | Yagaum | **0.62 (0.41, 0.92)** | **0.018** | **0.40 (0.27, 0.58)** | **<0.001** | **0.54 (0.36, 0.81)** | **0.003** | 0.78 (0.54, 1.13) | 0.182 | **0.43 (0.28, 0.65)** | **<0.001** |
|  | Alexishafen | 1.28 (0.84, 1.94) | 0.247 | 1.06 (0.72, 1.56) | 0.754 | **0.18 (0.11, 0.29)** | **<0.001** | **0.50 (0.35, 0.71)** | **<0.001** | **0.16 (0.10, 0.26)** | **<0.001** |
|  | Mugil | **0.43 (0.30, 0.62)** | **<0.001** | **0.54 (0.38, 0.77)** | **0.001** | 1.02 (0.72, 1.45) | 0.897 | **1.59 (1.13, 2.23)** | **0.007** | 1.09 (0.74, 1.58) | 0.671 |
| **Education level (years)** |  |  |  |  |  |  |  |  |  |  |  |
|  | 0-2 (Elementary) | Ref |  | Ref |  | Ref |  | Ref |  | Ref |  |
|  | 3-8 (Primary) | 0.95 (0.55, 1.63) | 0.852 | 1.11 (0.66, 1.85) | 0.705 | 1.61 (0.88, 2.96) | 0.125 | 1.05 (0.63, 1.77) | 0.853 | 1.34 (0.74, 2.42) | 0.329 |
|  | 9-12 (Secondary) | 1.04 (0.59, 1.83) | 0.900 | 1.31 (0.77, 2.25) | 0.321 | 1.82 (0.97, 3.41) | **0.061** | 1.18 (0.69, 2.03) | 0.541 | 1.34 (0.73, 2.47) | 0.418 |
|  | 13 (Tertiary) | 1.34 (0.59, 3.05) | 0.492 | **2.39 (1.08, 5.26)** | **0.031** | **2.65 (1.17, 5.99)** | **0.020** | 1.32 (0.63, 2.76) | 0.460 | 1.56 (0.69, 3.49) | 0.282 |
| **Employment status** |  |  |  |  |  |  |  |  |  |  |  |
|  | Unemployed/student | Ref |  | Ref |  | Ref |  | Ref |  | Ref |  |
|  | Informal | 1.08 (0.68, 1.71) | 0.743 | 1.07 (0.69, 1.67) | 0.758 | 1.05 (0.65, 1.68) | 0.851 | 0.91 (0.59, 1.90) | 0.657 | 0.98 (0.62, 1.57) | 0.942 |
|  | Formal | 0.97 (0.54, 1.73) | 0.911 | 0.96 (0.55, 1.65) | 0.870 | 0.79 (0.44, 1.42) | 0.434 | **0.59 (0.35, 0.99)** | **0.046** | 0.71 (0.40, 1.28) | 0.2576 |
| **Gravidity** |  |  |  |  |  |  |  |  |  |  |  |
|  | 1 | Ref |  | Ref |  | Ref |  | Ref |  | Ref |  |
|  | 2-3 | 1.22 (0.91, 1.64) | 0.189 | **1.28 (0.97, 1.69)** | **0.087** | 1.27 (0.93, 1.73) | 0.134 | 1.19 (0.90, 1.56) | 0.218 | 1.12 (0.83, 1.53) | 0.456 |
|  | ≥4 | 1.34 (0.93, 1.94) | 0.116 | **1.40 (0.99, 1.99)** | **0.057** | **1.70 (1.17, 2.49)** | **0.006** | **1.40 (0.99, 1.96)** | **0.054** | 1.23 (0.75, 1.55) | 0.671 |
| **Ever used alcohol** |  |  |  |  |  |  |  |  |  |  |  |
|  | No | Ref |  | Ref |  | Ref |  | Ref |  | Ref |  |
|  | Yes | **1.40 (0.97, 2.02)** | **0.075** | 1.10 (0.79, 1.54) | 0.579 | 1.08 (0.75, 1.55) | 0.688 | 0.93 (0.68, 1.29) | 0.672 | 1.07 (0.74, 1.54) | 0.716 |
| **Cooking method (open fire/coal)** |  |  |  |  |  |  |  |  |  |  |  |
|  | No | Ref |  | Ref |  | Ref |  | Ref |  | Ref |  |
|  | Yes | **0.50 (0.23, 1.08)** | **0.079** | 1.13 (0.60, 2.11) | 0.713 | 1.26 (0.68, 2.34) | 0.461 | 1.55 (0.86, 2.77) | 0.142 | 1.05 (0.18, 1.07) | 0.886 |

CI, confidence interval; OR, odds ratio.

#### How and why the study was conducted

In a multivariable regression analysis, study site was the only predictor associated with participants’ preference for receiving information on why and how the study was conducted (Table 3). Compared with women attending the urban Modilon/Town clinics, women attending the rural Yagaum (aOR 0.62, 95%CI 0.41, 0.92) and Mugil clinics (aOR 0.43, 95%CI 0.30, 0.62) were less likely to prefer this information.

#### Medical and scientific advances

Study site and education level were associated with participants’ preferences for receiving information on medical and scientific advances (Table 3). Compared with women attending the urban Modilon/Town clinics, women attending the rural Yagaum (aOR 0.40, 95%CI 0.27, 0.58) and Mugil clinics (aOR 0.54, 95%CI 0.38, 0.77) were less likely to prefer this information. By contrast, women with tertiary education were more likely to prefer information on medical and scientific advances than women with no formal or elementary education (aOR 2.39, 95%CI 1.08, 5.26).

#### Future research and policy implications

Study site, education level, and gravidity were associated with participants’ preference for receiving information on future research and policy changes (Table 3). Compared with women attending the urban Modilon/Town clinics, women attending the Alexishafen (aOR 0.18, 95%CI 0.11, 0.29) and Yagaum clinics (aOR 0.54, 95%CI 0.36, 0.81) were less likely to prefer this information. In contrast, women with tertiary education were more likely to prefer this content than women with no formal or elementary education (aOR 2.65, 95%CI 1.17, 5.99). Women with four or more pregnancies were also more likely to prefer this information than women in their first pregnancy (aOR 1.70, 95%CI 1.17, 2.49).

#### Individual study results

Study site and employment status were associated with participants’ preference for receiving their individual study results (Table 3). Compared with women attending the urban Modilon/Town clinics, women attending the peri-urban Alexishafen clinic were less likely to prefer receiving their individual study result (aOR 0.50, 95%CI 0.35, 0.71), whereas women attending the rural Mugil clinic were more likely to prefer this information (aOR 1.59, 95%CI 1.13, 2.23). Women in formal employment were less likely to prefer receiving their individual study results than unemployed women/students (aOR 0.59, 95%CI 0.35, 0.99).

#### Multiple preferences

Study site and gravidity were associated with participants selecting multiple content preferences (Table 3). Compared with women attending the urban Modilon/Town clinics, women attending the peri-urban Alexishafen (aOR 0.16, 95% CI 0.10, 0.26) and rural Yagaum clinics (aOR 0.43, 95%CI 0.28, 0.65) were less likely to select multiple content preferences.

## Discussion

Almost all participants wanted to learn about the results of the clinical trial they participated in. They viewed this as an acknowledgement of their contribution to research and an opportunity to fully understand the study results. Most of them preferred a written letter summary of the study results and a meeting at the nearest clinic for results dissemination. More than half of participants wanted to learn about study results when they are good, the purpose of the study, advances in treatment, and how the study was conducted. Around half of participants also wanted to receive their own individual study results or results specific to them.

Our findings reinforce the growing recognition that dissemination of study results is an important component of clinical research rather than an optional post-trial activity. Almost all participants for whom a questionnaire could be administered wished to receive study results, consistent with previous studies demonstrating that participants value dissemination as a sign of respect, transparency and reciprocity (8, 11, 19). Participants valued dissemination not only as an opportunity to understand the study in which they participated, but also as recognition of their contribution to the research (5, 20). The importance placed on acknowledgement of participation is consistent with findings from the recent multi-country malaria trial, where acknowledgement of trial participation was also the dominant motivation among participants in Pakistan, whereas Ethiopian participants placed greater emphasis on understanding the benefits of the research for their community (11). Together these results suggests that while dissemination is widely valued, the reasons participants value it are context specific(5). Acknowledgement of participation may be particularly important in settings where research relies heavily on long-term relationships between communities and research institutions (21).

While previous work has demonstrated differences in dissemination preferences between countries, our findings suggest that meaningful heterogeneity may also exist within countries and even between communities participating in the same clinical trial (11). Participants living in the catchment area of and attending the rural Mugil clinic were more likely to favour community-based dissemination, while those attending the peri-urban Alexishafen clinic more often preferred written materials, personalised communication, and multiple dissemination approaches. Preferences for the content of dissemination materials also varied between communities. Although this study was not designed to determine the reasons underlying these differences, they are likely to reflect broader contextual factors, such as differences in communication practices, previous research experience, or local health service organisation.(22)

Despite these local differences, overall participants showed a clear preference for low-technology dissemination approaches, particularly written summaries and community-based meetings, whereas electronic communication methods were rarely selected. Previous dissemination initiatives in low-and middle-income countries have shown that community-based approaches using face-to-face meetings and locally appropriate communication materials can successfully return study findings to participants and communities (11, 23). By contrast, patients in high-income settings often prefer electronic methods of results dissemination via emails, through a website or social media platforms, phone call or messages via messengers, or short message services (24).

One particularly important finding was that approximately half of participants wished to receive their own individual study results. This was substantially higher than reported in the recent multi-country malaria trial, where only around one-fifth of participants expressed this preference (11, 25). This suggest that in the PNG context, participants may place greater value on information directly related to their own health and study participation, in particular at rural sites. Whilst results of point-of-care tests, such as haemoglobin checks or ultrasound scans, were immediately communicated to patients and their caregivers and influenced their care, participants may also be interested in results of later laboratory analyses that were done as part of the trial. This has implication on study conduct and dissemination practice, and researchers should consider how participant expectations regarding individual results are managed during the consent process and throughout trial follow-up. However, returning individual study results is not always practical or scientifically appropriate. In large multi-centre trials or observational studies, individual findings may have limited clinical relevance or be difficult to interpret without appropriate clinical context. Previous studies suggest that participants are generally less interested in receiving individual research results when these have limited or no immediate implications for their own health (26). In contrast, interest in individual results is considerably greater when the findings are directly relevant to disease management or future health outcomes, such as in chronic conditions including diabetes mellitus (27). Going forward it is therefore important to prospectively define which individual results can be returned to participants and when they should be communicated. While some clinically relevant results are available during the trial, others derived from downstream laboratory analyses may only become available months after study completion. Clearly communicating these differences to participants from the outset is essential to ensure that expectations regarding the return of individual results are appropriately managed.

Our findings on participants preferences will be used to inform the design of planned dissemination activities and provide practical guidance for future clinical research in PNG. Although dissemination preferences differed between study sites, the differences reflected the relative importance of different approaches rather than completely distinct preferences (25). The dissemination strategy therefore combines the two most preferred dissemination methods—a written summary and a community-based meeting—and offers both approaches to all participating communities. A written letter, prepared in an accessible format and language, will be distributed to participants by community volunteers. The letter will both summarise the study and serve as an invitation to a group dissemination meeting at the participants’ local health facility, where the findings will be presented and discussed with participants.

This study comes with several limitations. Firstly, the questionnaire was based on limited previous work and was not informed by extensive qualitative work exploring participants experiences with and expectations for study result dissemination (11, 17). Additional qualitative work may have identified further dissemination preferences or contextual factors that were not captured by the survey. Second, dissemination preferences were assessed using a questionnaire and therefore reflect stated preferences rather than participants’ experience of receiving study results. Preferences may change once participants have experienced different dissemination approaches, and further evaluation of the planned dissemination activities will be important to determine whether the preferred approaches identified in this study translate into positive participant experiences. Third, although the study included participants from urban, peri-urban, and rural communities, participants were recruited from a single province. Preferences may differ in other provinces with different cultural contexts. Furthermore, the study was conducted among pregnant women participating in a malaria prevention trial. Dissemination preferences may differ among men, non-pregnant adults, children or participants in other types of clinical research, limiting the broader generalisability of the findings. Lastly, our multivariable analyses on factors that influence choices regarding mode and content of dissemination cannot, by design, demonstrate causality, and was exploratory without adjusting for multiple comparisons, which needs to be taken into consideration when interpretating analysis findings.

## Conclusion

Almost all participants wanted to receive the results of the clinical trial in which they had participated, reinforcing the ethical responsibility of researchers to return study findings to participants and their communities. However, preferences for both the methods of dissemination and the information participants wished to receive varied considerably between study sites, highlighting that a single dissemination strategy is unlikely to meet the needs of all participants. These findings provide practical guidance for the design of participant-centered dissemination strategies and support the incorporation of dissemination planning into the design of future clinical trials in Papua New Guinea and other low- and middle-income settings.

## Acknowledgements

We sincerely thank all the participants who participated, the PNG Institute of Medical Research and Menzies School Health Research team, SAPOT Trial team (clinical, community liaison, community volunteers, admin and data staff) and the in charges of the participating clinics [Mugil Health Center, Alexishafen Health Center, Town clinic, Madang Provincial Hospital (Modilon) antenatal clinic and Yagaum Rural Hospital]. We acknowledge Professor Stephen Rogerson for his continuous support to the team.

## Abbreviations

aOR: adjusted Odd Ratio
CI: confidence interval
n/N: number of participants
IPTp: intermittent preventative treatment in pregnancy
PNG: Papua New Guinea
PNGIMR: Papua New Guinea Institute of Medical Research
SAPOT: sulphadoxine-pyrimethamine plus dihydroartemisinin-piperaquine for the prevention of malaria infection and adverse pregnancy outcomes

## Authors contributions

*Conceptualization*: Alice Mengi, Kamala Thriemer, Holger W Unger.

*Data curation*: Alice Mengi, Hellen Mnjala, Grant Lee, Holger W Unger.

*Formal analysis*: Alice Mengi, Mary Bagita -Vangana, Kamala Thriemer, Holger W Unger.

*Funding acquisition*: Holger W Unger, Moses Laman, John W Bolnga

*Investigation*: Alice Mengi, Paula Tesine, Janeth Kulimbao, Jacobeth Mase, Lennie S Mal, Grant Lee

*Methodology:* Alice Mengi, Mary Bagita -Vangana, Kamala Thriemer, Holger W Unger.

*Project administration*: Alice Mengi, Paula Tesine, Helen Mjnala *Supervision*: Holger W Unger, Kamala Thriemer. Sarah Cassidy-Seyoum

*Writing – original draft*: Alice Mengi, Mary Bagita-Vangana,

*Writing – review & editing*: Alice Mengi, Mary Bagita-Vangana, Kamala Thriemer, Holger W Unger, Sarah Cassidy-Seyoum, Moses Laman, Paula Tesine, John W Bolnga, Maria Ome-Kaius, Janeth Kulimbao, Jacobeth Mase, Lennie S Mal, Helen Mnjala, Grant Lee,

## Data availability

The minimal data set and the accompanying code is available from the corresponding authors upon request, abiding by the data protection policy of the Papua New Guinea Institute of Medical Research.

## Declarations

The were no existing competing interest declared among the authors.

